# Deep Learning-based Differentiation of Drug-induced Liver Injury and Autoimmune Hepatitis: A Pathological and Computational Approach

**DOI:** 10.64898/2026.03.05.26347708

**Authors:** Azuki Shimizu, Kosuke Imamura, Kaori Yoshimura, Atsushi Tanaka, Makoto Sato, Kenichi Harada

**Author notes:** Correspondence (M.S.), (K.H.).

## Abstract

Drug-induced liver injury (DILI) is an acute inflammatory liver disease caused not only by prescription and over-the-counter medications but also by health foods and dietary supplements. Typically, DILI patients recover once the causative substance is identified and discontinued. In contrast, autoimmune hepatitis (AIH) results from the immune-mediated destruction of hepatocytes due to a breakdown of self-tolerance mechanisms. Patients presenting with acute-onset AIH often lack characteristic clinical features, such as autoantibodies, and require prompt steroid treatment to prevent progression to liver failure. Liver biopsy currently remains the gold standard to differentiate acute DILI from AIH; however, general pathologists face significant diagnostic challenges due to overlapping histopathological features. This study integrates pathology expertise with deep learning-based artificial intelligence (AI) to differentiate DILI from AIH using histopathological images. Our AI model demonstrates promising classification accuracy (Accuracy 74%, AUC 0.81). This paper presents a detailed pathological analysis alongside AI methods, discusses the current model performance and limitations, and proposes directions for future improvements.

## Introduction

Drug-induced liver injury (DILI) and autoimmune hepatitis (AIH) are clinically significant liver diseases characterized by acute hepatic inflammation. DILI arises following exposure to medications, dietary supplements, or herbal products and often resolves once the causative agent is withdrawn. However, delayed recognition can result in severe liver injury and even liver failure [1,2]. In contrast, AIH is caused by immune-mediated hepatocyte destruction due to a breakdown of self-tolerance. It can present with nonspecific symptoms and may progress rapidly without timely corticosteroid treatment [3,4]. Accurate differentiation between DILI and AIH is therefore essential, as each condition requires fundamentally distinct therapeutic management.

Pathologically, differentiating DILI from AIH is notably difficult because the two entities share histological overlaps, including portal and intralobular inflammatory cell infiltration, primarily lymphocytes and plasma cells, hepatocellular necrosis and apoptosis, and centrilobular necrosis [5,6]. This overlap can be particularly problematic during acute presentations, when serological markers such as autoantibodies may be absent or inconclusive. As a result, pathologists frequently encounter significant diagnostic uncertainty, which can meaningfully affect clinical decision-making.

Liver biopsy remains the gold standard for differentiating DILI from AIH, yet histopathological diagnosis is inherently limited by subjective interpretation, inter-observer variability, and dependence on the experience of individual pathologists. Subtle or early-stage changes may be overlooked, contributing further to diagnostic ambiguity. These limitations underscore the need for more objective, reproducible, and standardized diagnostic tools to complement conventional pathology.

Recent advances in artificial intelligence (AI), particularly deep learning, have transformed computational pathology by enabling highly accurate and reproducible image-based diagnostics [7,8]. Convolutional neural networks (CNNs) can learn complex morphological patterns directly from raw histological images, reducing human bias and enhancing consistency. AI models have demonstrated strong performance across cancer classification, prognosis prediction, and quantitative assessment of histological features, thereby streamlining pathological workflows and supporting pathologists in challenging cases. Extending this approach beyond oncology, AI has begun to show promise in inflammatory and autoimmune diseases, including liver disorders.

In this study, we developed a deep learning–based AI model specifically designed to distinguish DILI from AIH using multi-institutional histopathological data collected by expert hepatopathologists across Japan. By integrating explainability approaches, including Grad-CAM and Guided Backpropagation, we further evaluated the morphological basis of the model’s predictions. Through this combined analytical framework, our work aims not only to establish the quantitative and reproducible diagnostic tool for this difficult differential diagnosis, but also to uncover disease-relevant histological patterns that are not readily accessible through conventional pathology.

## Pathological Methods

### Sample collection and patient selection criteria

Formalin-fixed paraffin-embedded (FFPE) liver biopsy specimens were obtained from patients who had been clinically diagnosed or suspected with either DILI or AIH. Cases were selected retrospectively from institutional archives by board-certified pathologists. The diagnosis of DILI or AIH was made according to internationally accepted clinical and histological criteria, including clinical history, laboratory findings, and pathological evaluation. Cases with insufficient clinical information, inadequate tissue quality, or indeterminate pathological diagnosis were excluded from the study.

### Histological staining and slide preparation

Biopsy specimens were processed by routine histopathological methods. Sections of 3–5 μm thickness were cut from FFPE blocks and stained with hematoxylin and eosin (H&E) following standard protocols. Additional special stains (e.g., Masson’s trichrome or reticulin) were available for diagnostic support but were not used for the computational analyses in this study. The stained slides were scanned to generate whole-slide images using a NanoZoomer digital slide scanner (Hamamatsu Photonics, Hamamatsu, Japan) at ×20 magnifications. Digital images were exported in ndpi format for subsequent computational preprocessing and deep learning analysis.

## Computational Methods

### Image preprocessing

The original dataset consisted of a total of 196 images (ndpi format, Virtual Slide Scanner, HAMAMATSU, 442 nm/pixel), with 98 representing DILI and 98 representing AIH, were exported as jpg files using ndpitools (8bit RGB) [9], which were cropped into multiple 224×224 size images using a custom-made MATLAB code. Upon cropping, each pixel was classified into ‘tissue’ and ‘background’ using K-means clustering (Brighter pixels were regarded as ‘background’). If more than 99.9% of the cropped image were ‘background’, the image was omitted. While background pixels should be avoided as much as possible, the lumenal structure and the boundary between ‘tissue’ and ‘background’ may have important information. Therefore, the original image was divided to 5×5 pixel grids. If more than 10% of a grid is ‘tissue’, all pixels in the grid were regarded as ‘tissue’ expanding the tissue region. 224×224 size images were cropped avoiding the modified ‘background’ areas to obtain 125,685 DILI (90,800 training, 18,801 validations, and 16,084 test) and 160,095 AIH (118,214 training, 22,410 validations, and 19,471 test) images.

### AI model architecture

We utilized transfer learning based on GoogLeNet, a pretrained convolutional neural network originally trained on the ImageNet dataset. The final fully connected and classification layers of GoogLeNet were replaced to match the two output classes (DILI and AIH). Each image was resized to 224×224 pixels, in accordance with the input requirements of GoogLeNet. Images were augmented using random X/Y flipping and random X/Y translation (−10~+10) to increase dataset diversity and reduce overfitting. We evaluated the impact of stain normalization and color correction during preliminary experiments. However, applying stain normalization did not improve model performance and, in some settings, reduced classification accuracy. Therefore, no stain normalization or color correction was applied prior to training. To further justify model selection and assess architectural robustness, we additionally evaluated EfficientNet-B0.

### Data splitting and training procedures

All 196 samples were derived from independent patients. Whole-slide images were cropped as described above, and tiles originating from the same patient were stored within the same directory. Images were imported into MATLAB using *imageDatastore*. For each experimental setting (Fig. 1, S1, and S2), dataset partitioning was performed at the patient level using predefined index lists, ensuring that all tiles from a given patient were assigned exclusively to either the training, validation, or test set. The absence of overlap between training, validation, and test sets was explicitly confirmed using the *intersect* function.

**Fig. 1.**
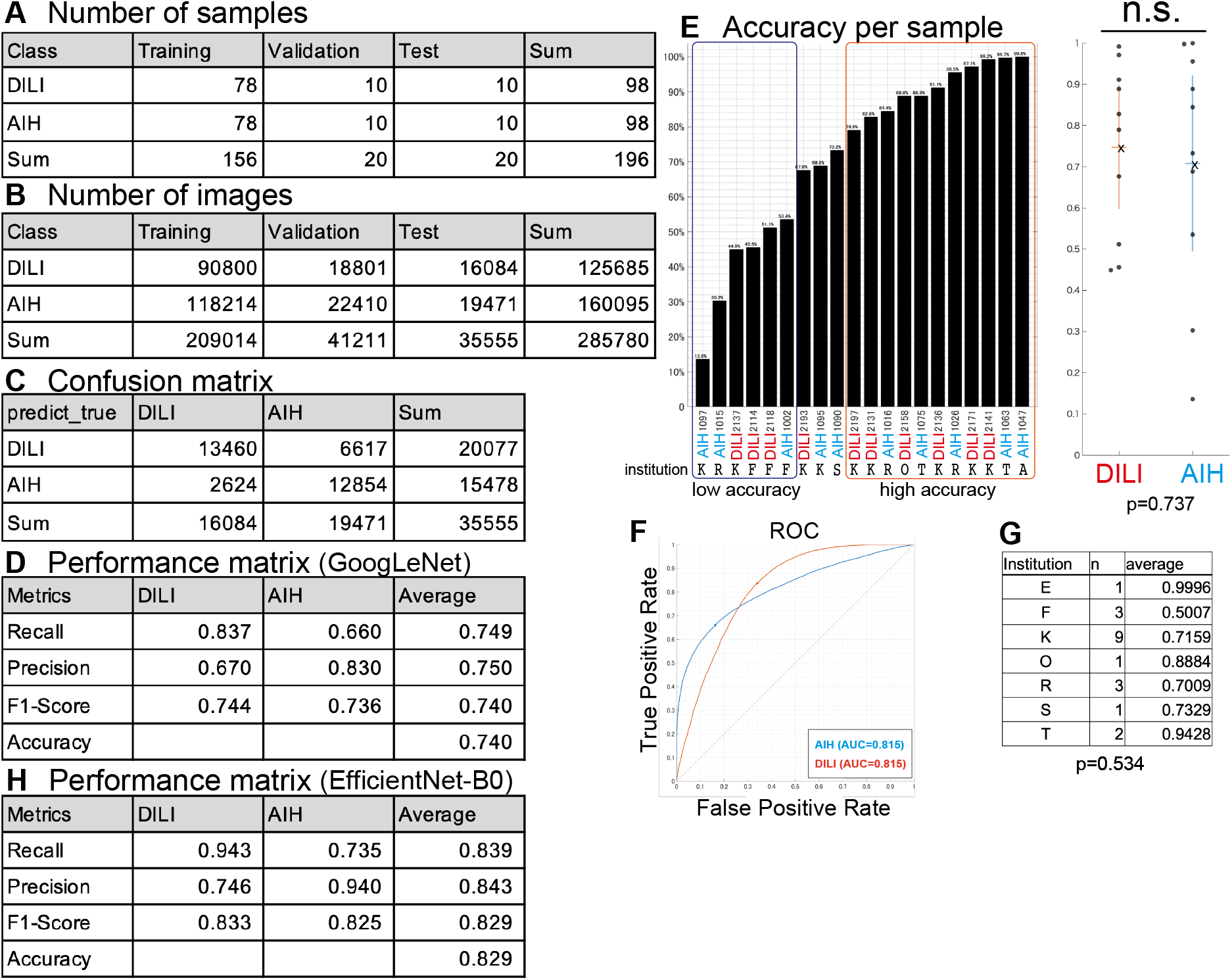
Dataset composition and overall performance of the CNN. (A) Number of samples used for training, validation, and testing. (B) Number of cropped images obtained from each sample for training, validation, and testing. (C) Confusion matrix of CNN predictions on the test dataset. (D) Performance metrics including recall, precision, F1-score, and accuracy. (E) Classification accuracy for each test sample. Black bars represent accuracy per sample, and the scatter plot shows the distribution of accuracies for DILI (red) and AIH (blue). No significant difference was observed between DILI and AIH samples (p=0.737). (F) Receiver operating characteristic (ROC) curves for DILI and AIH classification with operating points, showing an area under the curve (AUC) of 0.815 for both classes. (G) Average classification accuracy of samples from each institution. The number of test samples (n) and the mean accuracy are shown. One-way ANOVA revealed no significant difference among institutions (F(6, 13)=0.882, p=0.534). (H) Performance metrics based on an alternative model (EfficientNet-B0).

Training was conducted until the loss converged to a low and stable value using MATLAB’s Deep Learning Toolbox with GPU: NVIDIA GeForce RTX 3060 Ti and CPU: 13th Gen Intel Core i7-13700KF. The dataset was split into training, validation, and test sets in a ratio of about 80:10:10. The training parameters were set as follows. Solver: ADAM, MaxEpochs: 5, MiniBatchSize: 128, Shuffle: ‘every-epoch’, InitialLearnRate: 1.0000e-03, LearnRateSchedule: ‘none’, GradientDecayFactor: 0.9000, SquaredGradientDecayFactor: 0.9990, Epsilon: 1.0000e-08, L2Regularization: 1.0000e-04.

### Performance evaluation metrics (accuracy, sensitivity, specificity, ROC-AUC)

Model performance was evaluated using accuracy, precision, recall, F1-score, and confusion matrix. These metrics were calculated based on the test set results to assess the classification ability of the model in distinguishing between DILI and AIH. ROC-AUC was calculated and plotted using *rocmetrics* in MATLAB.

### Clustering the CNN evaluation

We analyzed the 1,024-dimensional output vectors of the global average pooling layer in the GoogLeNet CNN using PCA, UMAP, and t-SNE in Python. For the t-SNE analysis, we pre-compressed the original output vectors to 100-dimensional vectors using PCA and then compressed them further using t-SNE (perplexity = 100).

### Explainability analysis

Grad-CAM (Gradient-weighted Class Activation Mapping) and Guided Backpropagation were combined to create the heatmap showing the critical histological features for distinguishing AIH and DILI (Guided Grad-CAM) [10,11]. The *gradCAM* and *gradientAttribution* outputs in MATLAB were multiplied to create the heatmaps for AIH and DILI.

## Computational Results

### AI model classification performance summary

After transfer learning based on GoogLeNet using 90,800 training, 18,801 validation DILI images and 118,214 training, 22,410 validation AIH images, the obtained CNN was used to classify 16,084 DILI and 19,471 AIH test images showing 74.0% accuracy, 74.9% recall, 75.0% precision and 74.0% F1-score (Fig. 1A-D). The ROC indicates 0.815 AUC for DILI and AIH, suggesting that the CNN properly classifies DILI and AIH images (Fig. 1F). Two additional random splits of the dataset were analyzed and produced consistent results (Supplementary Figs. S1, S2). For each dataset partition used in Figs. 1, S1, and S2, EfficientNet-B0 was trained and tested under the same patient-level data splitting strategy and preprocessing conditions, yielding the consistent results (Figs. 1H, S1H, and S2H).

### Sample dependence of the classification performance

The results indicate that DILI and AIH are significantly correctly classified, but an accuracy of about 74% is not sufficient for actual diagnosis. Because pathology specimens are obtained independently at each hospital or research institute, staining conditions vary slightly. We investigated the extent to which the performance of AI-based classification depends on institutional and sample-specific variations.

Interestingly, classification accuracy was greater than 95% for some sample-derived images (Fig. 1E; sample #1047, 1063, 2141, 2171, 1026), while some samples yielded accuracy of less than 50% (Fig. 1E; sample #1097, 1015, 2137, 2114). We examined whether this difference in accuracy depended on whether the sample was a DILI or AIH, but no significant difference was obtained (Fig. 1E, p=0.737). Thus, we found that there are samples that can be classified very accurately and those that cannot, regardless of whether the sample is a DILI or AIH. Consistent results were obtained by two additional random splits of the dataset (Supplementary Figs. S1, S2).

### Institution-dependent differences in classification accuracy

Identifying the causes of sample-dependent differences in classification accuracy could improve classification performance. We analyzed the output of the global average pooling layer in the CNN (GoogLeNet) by compressing the 1,024-dimensional information to two dimensions using principal component analysis (PCA), uniform manifold approximation and projection (UMAP), and t-distributed stochastic neighbor embedding (t-SNE) (Fig. 2).

**Fig. 2.**
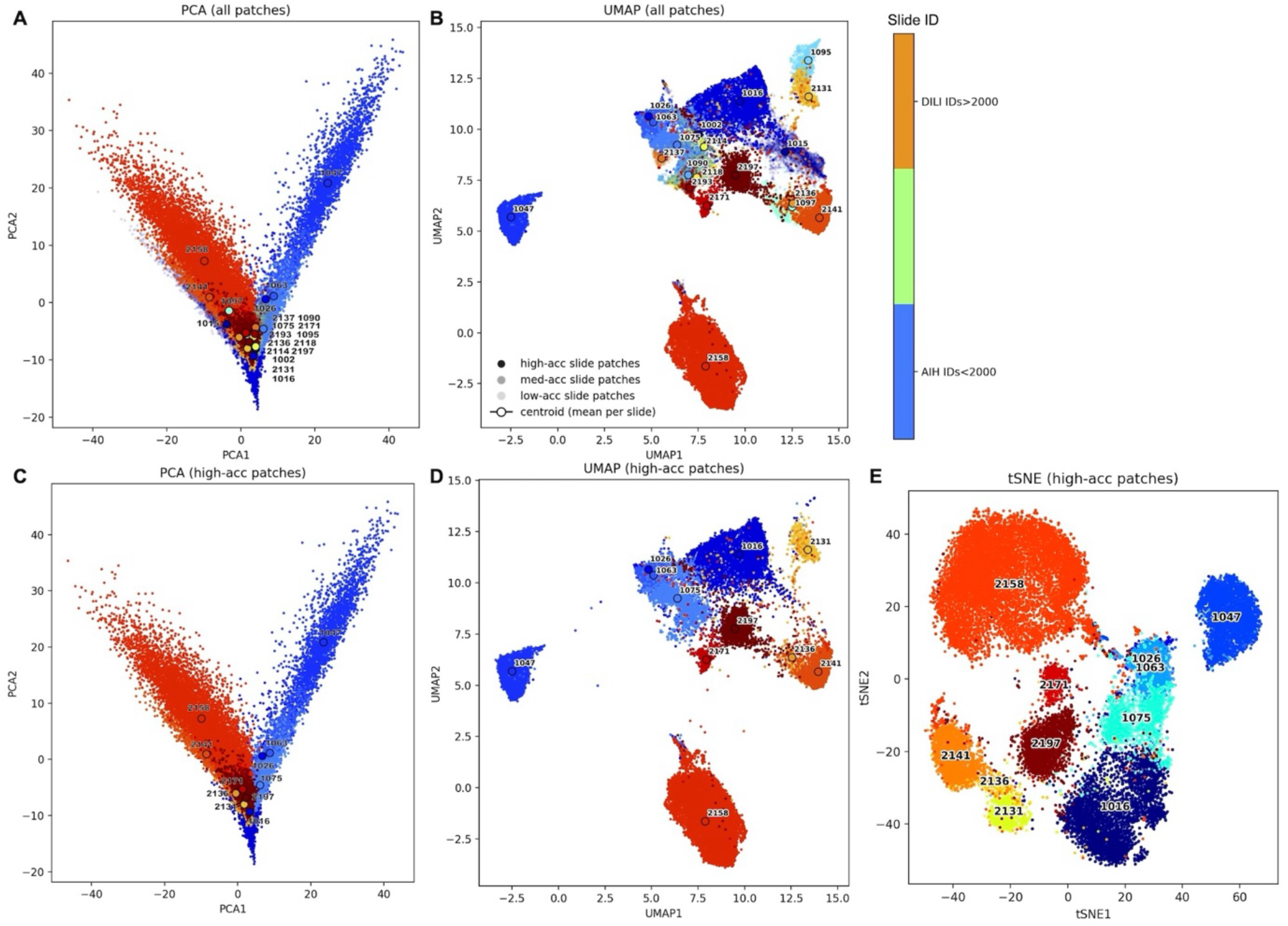
Clustering analyses of CNN feature representations and institutional differences in classification accuracy. (A, B) PCA and UMAP of all test samples (10 AIH and 10 DILI). Slide IDs <2000 represent AIH (blue), and slide IDs >2000 represent DILI (red). AIH and DILI clusters were intermingled and difficult to separate. (C, D) PCA and UMAP analyses of high-accuracy samples (accuracy >75%), showing clearer separation between AIH and DILI. (E) t-SNE analysis of high-accuracy samples, further demonstrating distinct clustering of AIH and DILI.

First, we analyzed all cropped images from the 20 test samples (10 AIH and 10 DILI) using PCA and UMAP (Fig. 2A, B). Slide IDs smaller than 2000 represent the AIH samples colored in blue tones, and slide IDs larger than 2000 represent the DILI samples colored in red tones. However, the AIH and DILI clusters were intermingled and difficult to separate in these analyses.

Next, we performed PCA and UMAP analyses focusing on samples with high accuracy (accuracy > 85%; Fig. 2C, D). Interestingly, we obtained separate clusters of AIH and DILI. Additionally, we performed a t-SNE analysis on the high-accuracy samples and observed a clear separation between the AIH and DILI clusters (Fig. 2E). These results suggest that the information embedded in the global average pooling layer is sufficient to distinguish between AIH and DILI in high-accuracy samples.

Unlike the high-accuracy samples, the low-accuracy samples formed significantly more intermingled clusters (Fig. 2A, B). We investigated whether the difference in accuracy was related to the institutions where the samples were prepared. The primary ANOVA analysis showed no statistical difference among institutions (Fig. 1G; ANOVA, F(6, 13)=0.882, p=0.534). Two additional random splits of the dataset yielded no consistent evidence of institutional effects (Supplementary Figs. S1, S2). One split suggested a mild difference (Fig. S1; ANOVA, F(4, 20)=3.085, p=0.039; F-critical=2.866), whereas the other did not (Fig. S2; ANOVA, F(3, 20)=2.026, p=0.143). Taken together, these results indicate that institution-dependent variation was minimal and unlikely to influence the overall conclusions. The variation in sample-dependent accuracy may be due to the diversity of patient conditions and the regional distribution of lesions.

### Explainability analysis of the neural network

It is difficult to evaluate how scientifically reliable the obtained results are because we do not know what information in the image is used to make the decision. We performed the explainability analysis by applying Grad-CAM and Guided Backpropagation to the CNN obtained in the above (see Method section) [10,11].

Here, we focused specifically on classifying samples that were accurately classified (e.g., #1047 AIH and #2141 DILI; Fig. 3). The Grad-CAM heatmaps show a broader activation pattern compared to Guided Backpropagation and differ greatly between the AIH and DILI channels. In AIH samples, only the former is activated; in DILI samples, only the latter is activated. Conversely, the heatmaps of Guided Backpropagation are largely the same between the AIH and DILI channels. The Guided Grad-CAM is the product of multiplication of the Grad-CAM and Guided Backpropagation, and its result also show high specificity to AIH and DILI.

**Fig. 3.**
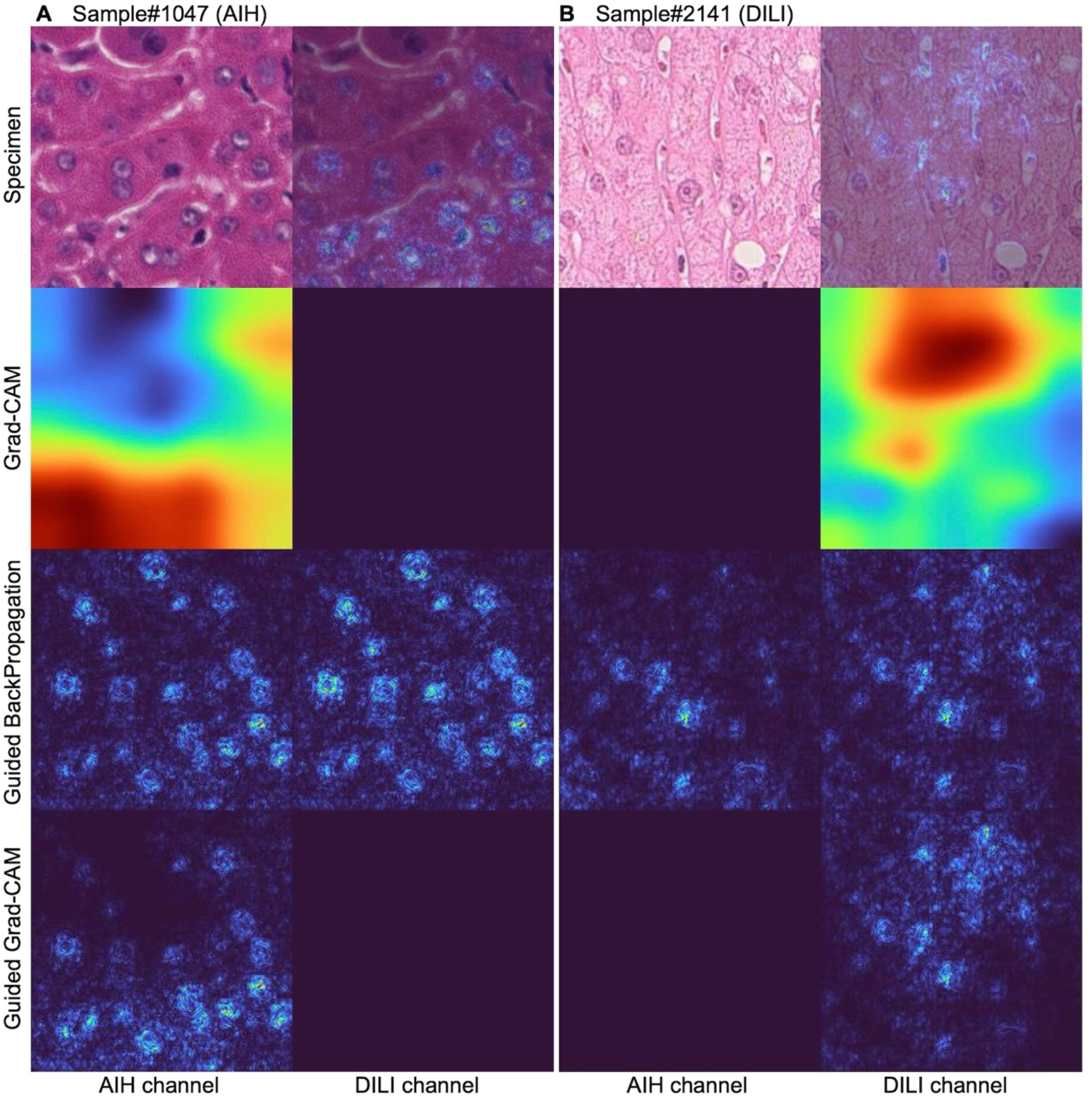
Explainability analysis of CNN-based classification using Grad-CAM and Guided Backpropagation. Representative (A) AIH (sample #1047) and (B) DILI (sample #2141) cases are shown. For each sample, the left column corresponds to the AIH channel and the right column to the DILI channel. Top row: H&E-stained images. The right-side images additionally display the Guided Grad-CAM results overlaid on the original H&E (AIH channel for the AIH sample, DILI channel for the DILI sample). Second row: Grad-CAM heatmaps highlighting global tissue regions contributing to classification. Third row: Guided Backpropagation heatmaps emphasizing pixel-level and nuclear morphological features. Bottom row: Guided Grad-CAM, obtained by element-wise multiplication of Grad-CAM and Guided Backpropagation, demonstrating high specificity to disease-related features.

Based on the results of Guided Backpropagation, the CNN appears to focus on nuclear morphology at the pixel level, although this alone is insufficient for distinguishing AIH from DILI. In contrast, Grad-CAM suggests that global tissue information about the tissue plays a significant role in the decision-making process.

## Pathological Results

### Description of distinctive histological features observed in DILI and AIH

Histological examination of liver biopsy specimens revealed overlapping but partly distinctive features between DILI and AIH.

In DILI, hepatocellular injury was often characterized by variable patterns of necrosis, including spotty and confluent necrosis, accompanied by mixed inflammatory infiltrates. The inflammatory infiltrates typically included lymphocytes, and eosinophils, with occasional cholestasis. The distribution and extent of injury varied across cases, reflecting the heterogeneous etiologies of DILI. In AIH, interface hepatitis was more consistently observed at a various intensity, with lymphoplasmacytic infiltrates extending into the periportal parenchyma. Prominent plasma cell infiltration was a characteristic feature. Hepatocyte rosettes and emperipolesis were mostly identified, supporting the diagnosis of AIH.

Although these features provide general diagnostic guidance [12,13], significant histological overlap between DILI and AIH was evident in many cases. This overlap highlights the inherent difficulty in distinguishing between the two entities based on histology alone. Computational analysis with CNNs, particularly guided Grad-CAM visualization (Fig. 3), suggested that tissue-level architecture (as highlighted by Grad-CAM) and nuclear morphology (as highlighted by Guided Backpropagation) may contribute differentially to disease classification.

## Discussion

### Clinical implications and importance of AI-assisted diagnostics

The differentiation between DILI and AIH remains a critical clinical challenge due to their overlapping histological presentations and divergent treatment strategies. Recent studies have demonstrated that artificial intelligence can be successfully applied to liver histology: for example, machine learning approaches have enabled quantitative measurement and monitoring of disease activity in NASH [14], and deep learning-based methods have been developed for the assessment of NAFLD/NASH progression [15].

Building on these advances, our study demonstrates that CNNs can achieve clinically relevant accuracy in distinguishing DILI from AIH based on histopathological images (Figs. 1, S1, S2). To the best of our knowledge, this is the first AI-based attempt to quantitatively differentiate DILI from AIH using multi-institutional biopsy data. Given the diagnostic difficulty and the absence of established computational approaches in this area, our results provide an important proof of concept.

Importantly, explainability analyses such as Guided Grad-CAM provided insights into the morphological features contributing to classification, suggesting that AI may serve not only as a diagnostic adjunct but also as a tool to highlight disease-relevant histological patterns (Fig. 3). The combination of deep learning with explainability further enabled us to extract pathological cues that are not readily captured by conventional visual inspection, offering new hypotheses regarding tissue-level and nuclear-level features relevant to differential diagnosis. Integrating AI into routine pathology workflows could support pathologists in challenging cases, reduce diagnostic variability, and ultimately contribute to timely therapeutic decisions.

### Limitations of pathology interpretations and computational model

Despite the promising performance of the proposed model, several limitations should be acknowledged. First, the pathological overlap between DILI and AIH remains substantial, and even expert pathologists may encounter difficulties in achieving consensus diagnoses. Although definitive diagnoses were established based on clinical follow-up, the intrinsic histopathological similarity between these conditions poses a fundamental diagnostic challenge.

Second, while 196 patient-derived whole-slide images were analyzed, this sample size remains modest for developing and validating a deep learning model for clinical application. The dataset was collected retrospectively from a limited number of institutions. Differences in tissue preparation, staining protocols, and slide-scanning procedures may influence model performance when applied to external cohorts. Although ANOVA analysis did not reveal a consistent institutional effect on model accuracy, the relatively small number of samples per institution limits the strength of this conclusion. Therefore, further validation using larger, multi-center datasets will be necessary to assess generalizability.

Interestingly, classification accuracy varied markedly among individual samples: some cases achieved accuracy exceeding 95%, whereas others fell below 50% (Figs. 1, S1, S2). These differences were independent of diagnostic category and institutional origin. This sample-dependent variability may reflect underlying biological heterogeneity, including differences in immune response, drug-induced injury mechanisms, inflammatory intensity, and lesion distribution within biopsy specimens. While further validation is needed, the combination of sample-level accuracy analysis and explainability tools highlights both the potential strengths of the model and important opportunities for refinement in future investigations.

### Strategies for performance enhancement

Building on the insights obtained in this study, several directions could further enhance the performance and clinical applicability of AI-assisted differentiation of DILI and AIH. First, expanding the dataset to include a larger number of well-annotated cases from diverse institutions will be essential to improve model robustness and generalizability. Our findings of marked sample-dependent variability highlight the importance of capturing the full clinical and histological heterogeneity of hepatitis cases. Increasing case diversity—not only in staining protocols but also in disease stage, lesion distribution, and patient background—would allow the model to learn broader and more representative features.

Standardizing staining and scanning protocols across institutions may further reduce technical variability and facilitate more consistent model training. From a computational standpoint, evaluating more advanced architectures, or vision transformers, as well as exploring ensemble approaches, has the potential to improve classification accuracy and stability beyond what was achieved with the current GoogLeNet-based model.

In addition, integrating multimodal data—including clinical parameters, laboratory findings, and serological markers—could help overcome limitations inherent to morphology-only diagnostics and may enable more precise disease stratification. The success of such integrative approaches will depend on continued collaboration between pathologists, hepatologists, and computational scientists. Ultimately, the insights from this study provide a foundation for building more powerful, interpretable, and clinically actionable AI-based diagnostic tools for challenging liver diseases.

## Acknowledgements

This work was supported by Grant-in-Aid for Scientific Research (B) and (C), Grant-in-Aid for Transformative Research Areas (A), from MEXT (22H05169, 22H05621, 24H00188, 24H01396, and 25K02282 to M.S., 23K06460 to K.H., and 24K14991 to K.I.).

## Author contributions

A.S. performed image preprocessing, CNN training, evaluation, and explainability analyses. M.S. conceived and designed the study, supervised the computational analyses, performed dimensionality-reduction analyses, prepared figures, and wrote the manuscript. K.H., K.Y, A.T. collected and curated the nationwide pathological dataset, provided pathological interpretation, and wrote the manuscript. K.I. advised on computational methodology and image-analysis strategies. All authors discussed the results, contributed to manuscript revision, and approved the final version.

## Data availability statement

The original whole-slide image data are derived from human patients and are not publicly available due to ethical and privacy restrictions. De-identified data may be made available from the corresponding author upon reasonable request and subject to approval by the institutional ethics committee.

The custom code newly generated in this study are available online (https://github.com/satouma7/Hepatitis).

## Ethics statement

This study was approved by the Kanazawa University Ethics Committee (Approval No. 2016-317 (1985)). All procedures were conducted in accordance with institutional guidelines and the Declaration of Helsinki. Patient data and whole-slide images were anonymized prior to analysis. Because this study was conducted retrospectively, individual informed consent was waived, and an opt-out consent process was implemented in accordance with institutional guidelines.

## Declaration of generative AI and AI-assisted technologies in the manuscript preparation process

During the preparation of this work the author(s) used ChatGPT, Gemini, and DeepL in order to write the code for the data analysis and draft the manuscript. After using this tool/service, the author(s) reviewed and edited the content as needed and take(s) full responsibility for the content of the published article.

## Figures

**Fig. S1.**
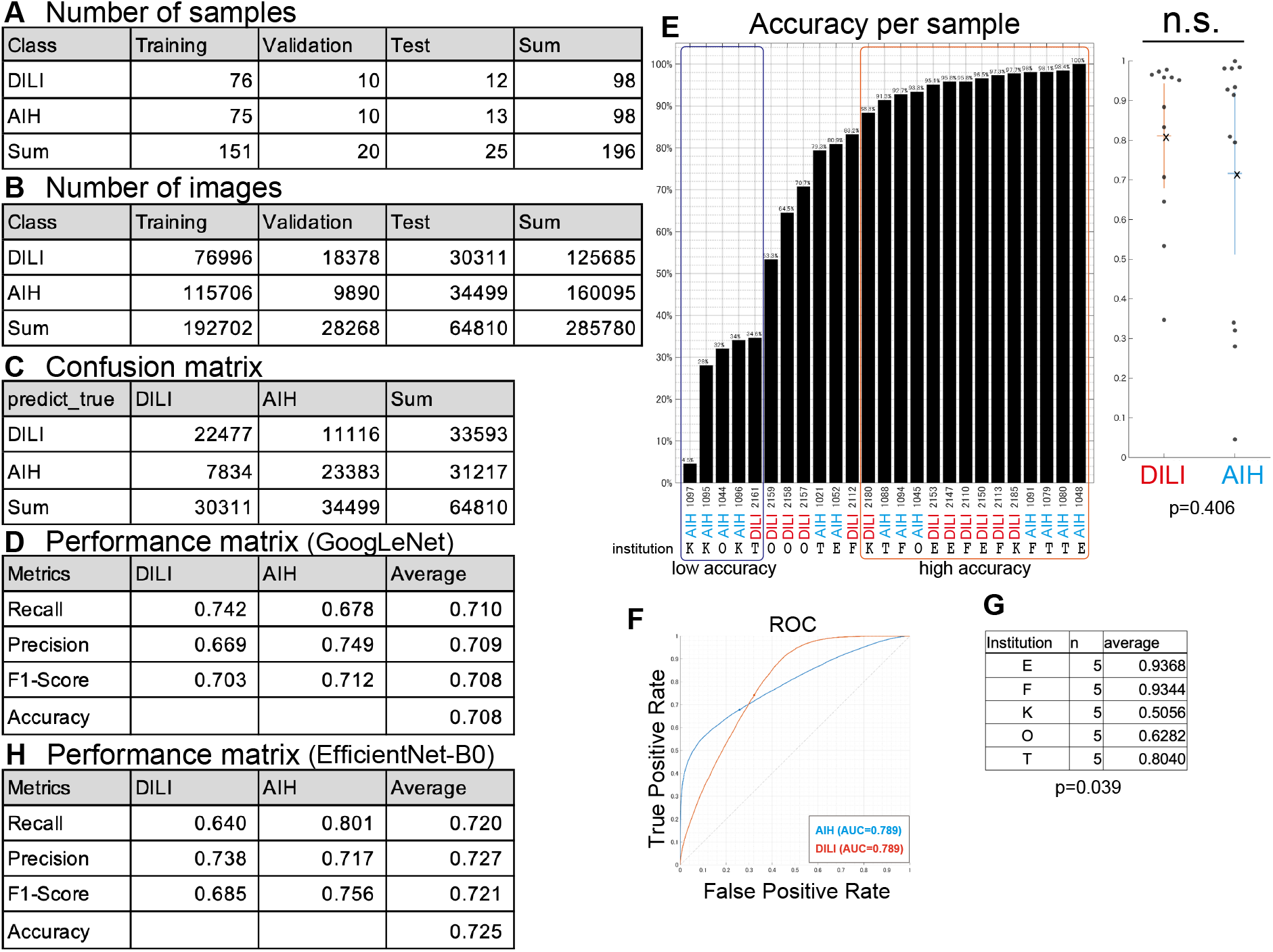
Additional random splits of the dataset 1. (A) Number of samples used for training, validation, and testing. (B) Number of cropped images obtained from each sample for training, validation, and testing. (C) Confusion matrix of CNN predictions on the test dataset. (D) Performance metrics including recall, precision, F1-score, and accuracy. (E) Classification accuracy for each test sample. Black bars represent accuracy per sample, and the scatter plot shows the distribution of accuracies for DILI (red) and AIH (blue). No significant difference was observed between DILI and AIH samples (p=0.406). (F) Receiver operating characteristic (ROC) curves for DILI and AIH classification with operating points, showing an area under the curve (AUC) of 0.789 for both classes. (G) Average classification accuracy of samples from each institution. The number of test samples (n) and the mean accuracy are shown. One-way ANOVA revealed no significant difference among institutions (F(4, 20)=3.085, p=0.039; F-critical=2.866). (H) Performance metrics based on an alternative model (EfficientNet-B0).

**Fig. S2.**
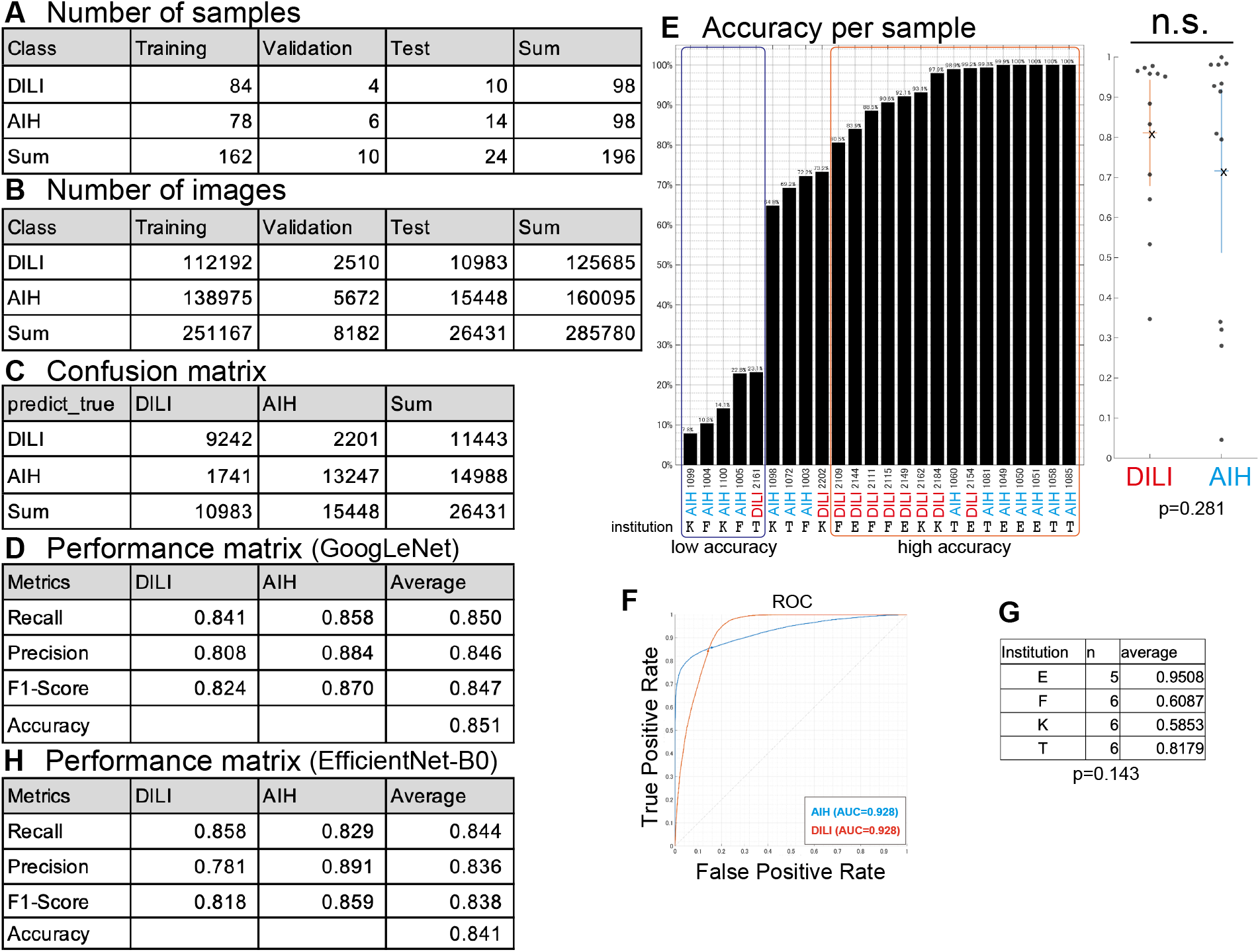
Additional random splits of the dataset 2. (A) Number of samples used for training, validation, and testing. (B) Number of cropped images obtained from each sample for training, validation, and testing. (C) Confusion matrix of CNN predictions on the test dataset. (D) Performance metrics including recall, precision, F1-score, and accuracy. (E) Classification accuracy for each test sample. Black bars represent accuracy per sample, and the scatter plot shows the distribution of accuracies for DILI (red) and AIH (blue). No significant difference was observed between DILI and AIH samples (p=0.281). (F) Receiver operating characteristic (ROC) curves for DILI and AIH classification with operating points, showing an area under the curve (AUC) of 0.789 for both classes. (G) Average classification accuracy of samples from each institution. The number of test samples (n) and the mean accuracy are shown. One-way ANOVA revealed no significant difference among institutions (F(3, 20)=2.026, p=0.143). (H) Performance metrics based on an alternative model (EfficientNet-B0).

